# Evidence of Tenofovir Resistance in Chronic Hepatitis B Virus (HBV) Infection: An Observational Case Series of South African Adults

**DOI:** 10.1101/2020.03.18.20038216

**Authors:** Jolynne Mokaya, Tongai G Maponga, Anna L McNaughton, Marije Van Schalkwyk, Susan Hugo, Joshua B Singer, Vattipally B Sreenu, David Bonsall, Mariateresa de Cesare, Monique Andersson, Shiraaz Gabriel, Jantje Taljaard, Eleanor Barnes, Wolfgang Preiser, Christo Van Rensburg, Philippa C Matthews

## Abstract

**Introduction:** Tenofovir disoproxil fumarate (TDF) is widely recommended for treatment of chronic hepatitis B virus (HBV) infection because it is safe, affordable and has a high genetic barrier to resistance. TDF resistance associated mutations (RAMs) have been reported, but data are limited, particularly for Africa. We set out to identify RAMs in individuals with detectable HBV viraemia on TDF treatment.

**Methods:** We recruited adults with chronic HBV infection from Cape Town, South Africa, identifying individuals with a TDF resistance phenotype, defined as persistent HBV vireamia despite >12 months of TDF treatment. We sequenced HBV DNA using MiSeq Illumina with whole genome target enrichment, and analysed to determine the genotype and identify potential TDF RAMs, based on a pre-defined list of polymorphisms.

**Results:** Among 66 individuals with chronic HBV, we identified three meeting our phenotypic definition for TDF resistance, of whom two were coinfected with HIV. The sequences grouped as genotypes A1 and D3. In one participant, the consensus HBV sequence had ten polymorphisms that have been described in association with TDF resistance. Significant treatment non-adherence in this individual was unlikely, as HIV RNA was suppressed. TDF RAMs were also present in HBV sequences from the other two participants, but other factors including treatment non-adherence may also have had a role in failure of HBV DNA suppression in these cases.

**Discussion:** Our findings add to the evidence that RAMs in HBV RT can underpin a TDF resistant phenotype. This is the first time these RAMs have been reported from Africa in association with clinical evidence of TDF resistance.

**Contribution to the Field Statement:** Treatment of chronic hepatitis B virus (HBV) infection with nucleos(t)ide analogues (NAs) is one of the key strategies that needs to be upscaled in order to achieve the 2030 United Nations elimination target for viral hepatitis. Tenofovir disoproxil fumarate (TDF) is widely recommended for the treatment of chronic HBV infection because it has a high genetic barrier to resistance. However, TDF resistance associated mutations (RAMs) have been reported, but data are limited, with a need for further investigation. Within a cross-sectional cohort of adults with chronic HBV infection recruited in Cape Town, South Africa, we describe combinations of HBV polymorphisms in three adults with detectable HBV viraemia whilst on TDF treatment. This is the first evidence of potential TDF resistance in adults being treated for chronic HBV in Africa and it adds to the growing evidence of TDF resistance globally. It remains necessary to advocate for the development of new antiviral treatments for chronic HBV infection if we are to attain elimination targets.

## INTRODUCTION

Tenofovir disoproxil fumarate (TDF) is a first line nucleoside analogue (NA) agent used to treat chronic hepatitis B virus (HBV) infection (1,2). TDF has a strong track record of safety and tolerability, is affordable and widely available, and successfully suppresses plasma viraemia below the limits of detection in the majority of patients taking treatment (2,3). In contrast to some other first line antiviral agents, the genetic barrier to TDF resistance is high (1,2). However, recent reports suggest the possible emergence of TDF resistance associated mutations (RAMs) in HBV, raising concerns about the possibility of a drug resistant phenotype in which patients remain viraemic on therapy (e.g. (4,5)). A comprehensive review of potentially relevant TDF RAMs is now available, but this report highlights significant gaps and limitations in the current literature, highlighting the need for further investigation (6).

In addition to its use in HBV, TDF is widely used as part of the backbone of antiretroviral therapy (ART) for HIV (7,8). Thus, in settings where HIV is widespread, population exposure to TDF is high, raising the potential risk of selection of resistance in both HIV and HBV (8). Monitoring HIV viraemia in coinfected patients receiving TDF-based regimens provides insights about treatment compliance, as suppression of HIV viraemia is a helpful surrogate for good adherence to therapy.

International sustainable development goals set out the target for elimination of viral hepatitis infections as a public health threat by the year 2030 (9). Upscaling treatment of chronic HBV infection will be a fundamental way to reduce morbidity, mortality and incidence (10,11). Pursuit of a better understanding of the prevalence and clinical significance of HBV RAMs is therefore essential; selection and dissemination of TDF resistant HBV would not only limit the success of antiviral therapy for individual patients, but would also represent a population-level threat, by undermining efforts to curb transmission (8).

We identified patients from within a cross-sectional cohort of adults with chronic HBV infection recruited in Cape Town, South Africa, in whom concerns about drug resistance were raised on account of persistent HBV viraemia despite TDF therapy. Sequencing HBV from these individuals confirmed the presence of mutations that have previously been described in association with TDF resistance.

## METHODS

### Clinical cohort

Adults were enrolled into a cross-sectional cohort of chronic HBV infection at Tygerberg Hospital, Cape Town, (‘OxSAHep’ HBV study), as previously described (12). All participants provided valid written informed consent in accordance with the Declaration of Helsinki. Ethics approval was provided by the University of Oxford Tropical Research Ethics Committee (ref. OXTREC 01-18) and Stellenbosch University Human Research Ethics Committee (HREC ref. N17/01/013). We identified cases with chronic HBV infection in whom HBV viraemia was ≥20,000 IU/ml despite being on TDF treatment for >12 months (Fig 1).

**Figure 1:**
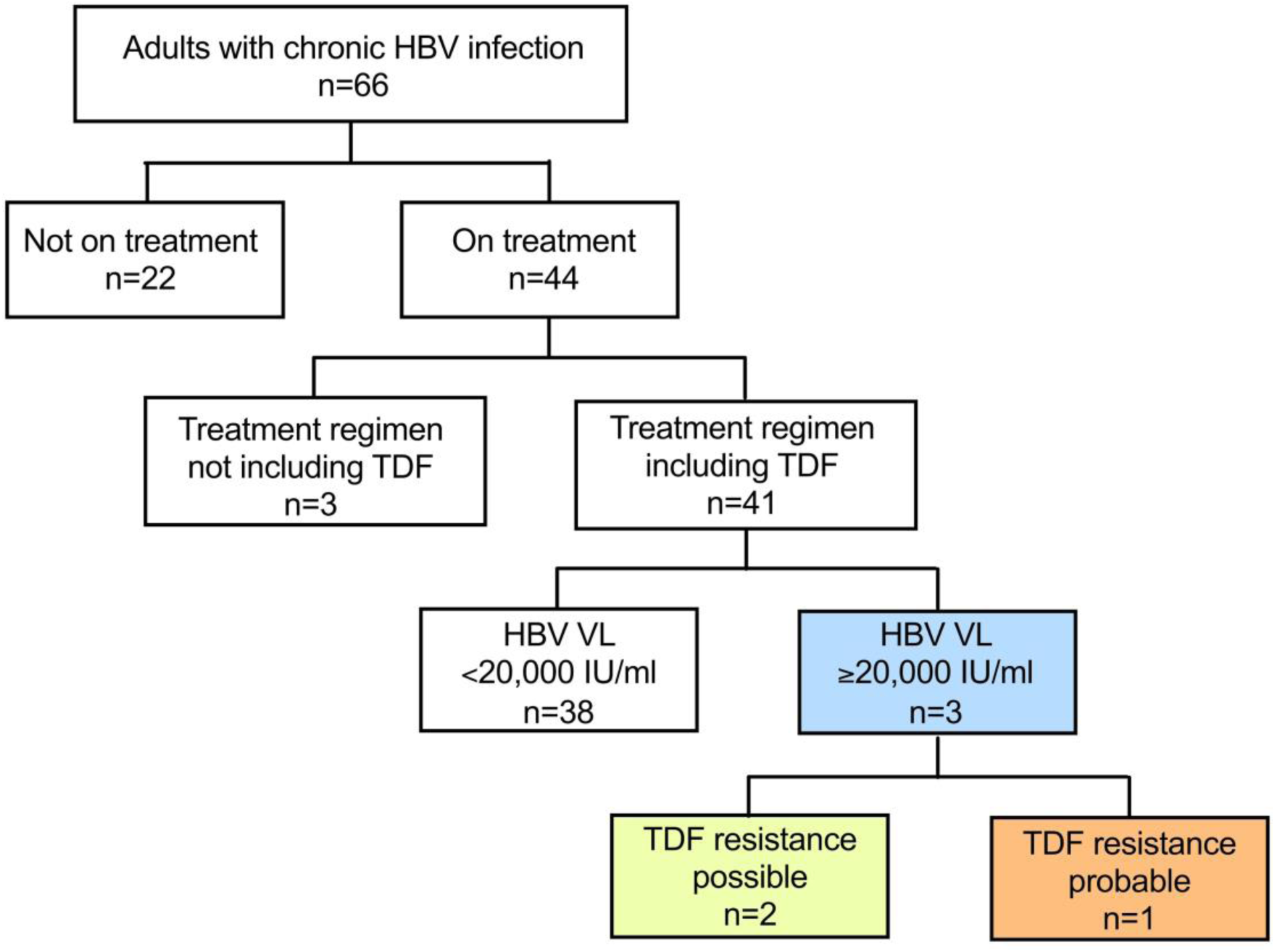
Selection of participants with chronic HBV infection for viral sequencing to identify tenofovir disoproxil fumarate (TDF) resistance associated mutations (RAMs). Adults attending clinical follow-up in Cape Town, South Africa, were enrolled over a period of one year. Three adults with persistent viraemia despite >12 months of TDF therapy are indicated in the blue box. Of these one had a phenotypic profile and HBV sequence data to support TDF resistance (orange box); two others had possible TDF resistance RAMs in HBV sequences but other factors including non-adherence to therapy may have contributed to persistent viraemia (yellow box).

### Antiviral therapy regimens

In this setting, patients are stratified for HBV therapy based on an algorithm that incorporates age, race, ALT, HBV DNA levels, imaging or biopsy evidence of fibrosis/cirrhosis, co-factors for liver disease, and family history. South African guidelines recommend use of lamivudine (3TC), entecavir (ETV) or tenofovir disoproxil fumarate (TDF) (13,14). In practice, TDF is most frequently used, as 3TC use is limited by concerns about drug resistance, and ETV is expensive and not widely available. TDF is a first-line component of the NRTI backbone of first-line combined antiretroviral therapy (ART) for HIV, and is also a frequent choice in second-line regimens (15,16), and for HIV infection TDF is often combined with emtricitabine (FTC) (13,14).

### Deep sequencing of HBV

HBV DNA was extracted from 0.5ml plasma samples using a NucliSENS magnetic extraction system (BioMérieux). HBV genomes were converted into fully double stranded (ds)DNA genomes using a completion-ligation approach based on a previously published protocol (17,18). DNA was purified with Agencourt RNAClean XP beads (Beckman Coulter) and libraries prepared with an NEBNext Ultra II FS DNA Library Prep Kit (New England Biolabs). A modified target-enrichment approach, based on the SeqCap EZ (Roche) protocol was used, with custom-designed HBV probes ordered from IDT (xGen Lockdown Probes). We sequenced the samples on an Illumina Mi-Seq using a v3 300bp paired end kit. Bioinformatic analysis was performed using Tanoti v1.3 (https://bioinformatics.cvr.ac.uk/software/tanoti/), with genotyping and variant using the HBV-GLUE resource (http://hbv-glue.cvr.gla.ac.uk/), based on the GLUE system (19).

### Identification of RAMs

We looked for mutations associated with TDF resistance using a catalogue of 37 different sites of polymorphism in HBV RT assimilated through systematic literature reviews (6,19). In order to identify polymorphisms that occur at consensus level in widely reported sequences, we downloaded all available full length sequences for Genotype A and D (n=840 and n=958, respectively) from HBVdb on the 20th November 2018.

## RESULTS

### Phenotypic evidence of TDF resistance

Among 66 adults with chronic HBV infection, we identified three with HBV DNA viral load ≥20,000 IU/ml in serum, despite therapy with TDF for ≥12 months (Fig 1), of whom two were co-infected with HIV. We used this level of persistent HBV viraemia on therapy as a phenotypic marker of patients with potential TDF-resistant HBV infection.

However, as an alternative to drug resistance, another explanation for persistent HBV viraemia on therapy is suboptimal adherence to a daily drug regimen. In two cases, we were able to use HIV viral load as a surrogate marker of adherence to therapy. In patient 209, HIV RNA suppression provides good evidence for adequate adherence to antiviral therapy (Fig 2A), increasing the probability that HBV drug resistance is genuinely present. In contrast, in patient 258, both HIV and HBV viraemia remained detectable, suggesting a possible impact of incomplete adherence to therapy, with or without drug resistance in one or both viruses (Fig 2B).

**Figure 2:**
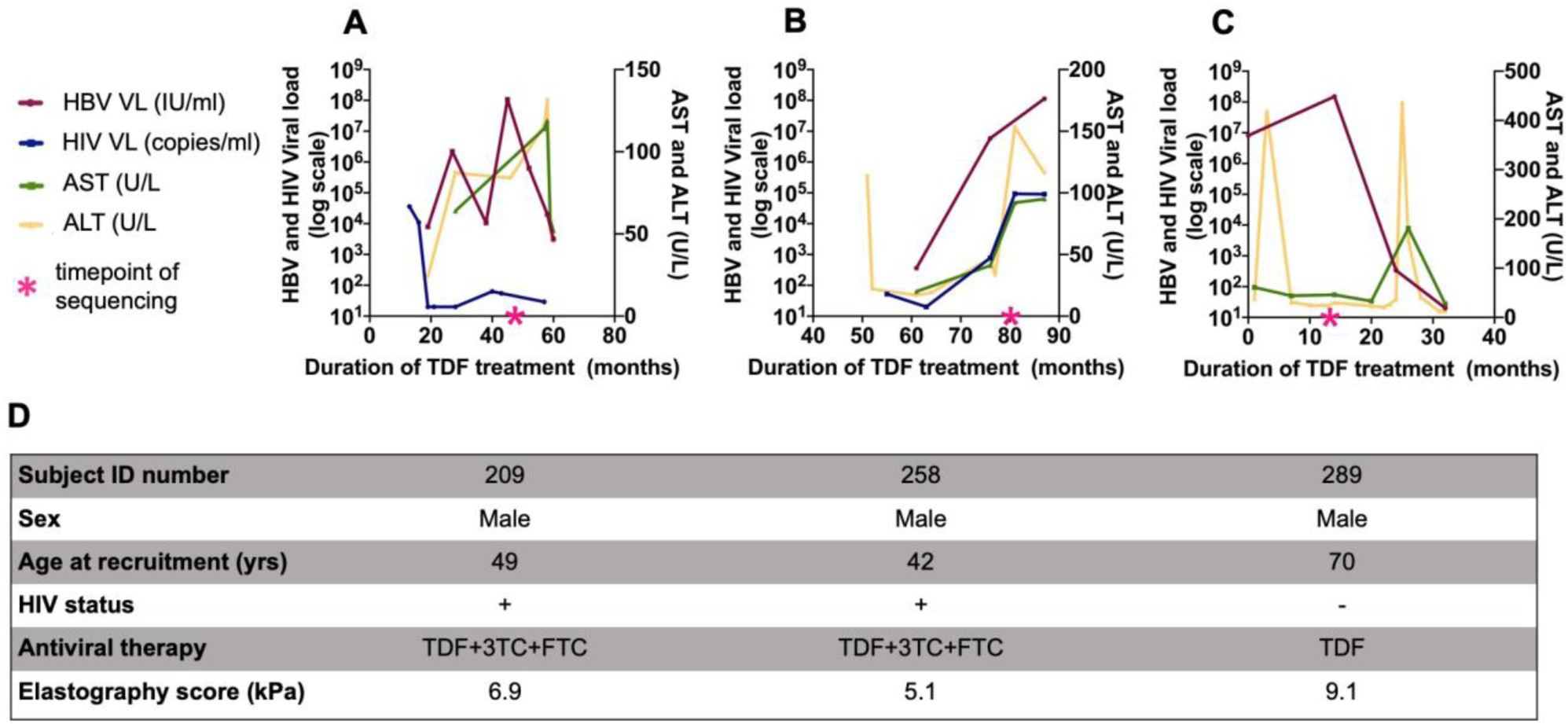
Longitudinal viral load and liver function test distribution of three adults with detectable HBV viraemia >20,000 IU/ml despite ≥12 months prescribed TDF therapy. Panels A-C plot laboratory parameters over time in three patients. The corresponding clinical data for each patient is shown below in panel D.

A third case (ID 289) was HIV negative. HBV viral suppression in this case eventually occurred after a cumulative 30 months on treatment (Fig 2C). It is recognised that HBV suppression may take many months of therapy even when the virus is deemed fully drug susceptible (20). Slow virologic suppression may be a better explanation of the clinical phenotype observed in this case, despite the potential RAMs identified at the time point sequenced. These explanations are not mutually exclusive, and may both have a role in explaining the time-delay to aviraemia, with or without a contribution of non-adherence to therapy.

These three patients reflect different phenotypes, illustrating how clinical observations and laboratory investigations can help to identify the presence of underlying viral drug resistance.

### Genotypic evidence of TDF resistance

We derived full length HBV genome sequences for all three patients, although with varying depth of coverage, identifying infection with genotype-A1 in two cases and -D3 in the third (Table 1). The HBV consensus sequences all contained between five and ten RAMs that have been previously described in association with TDF resistance (Table 1).

**Table 1:**
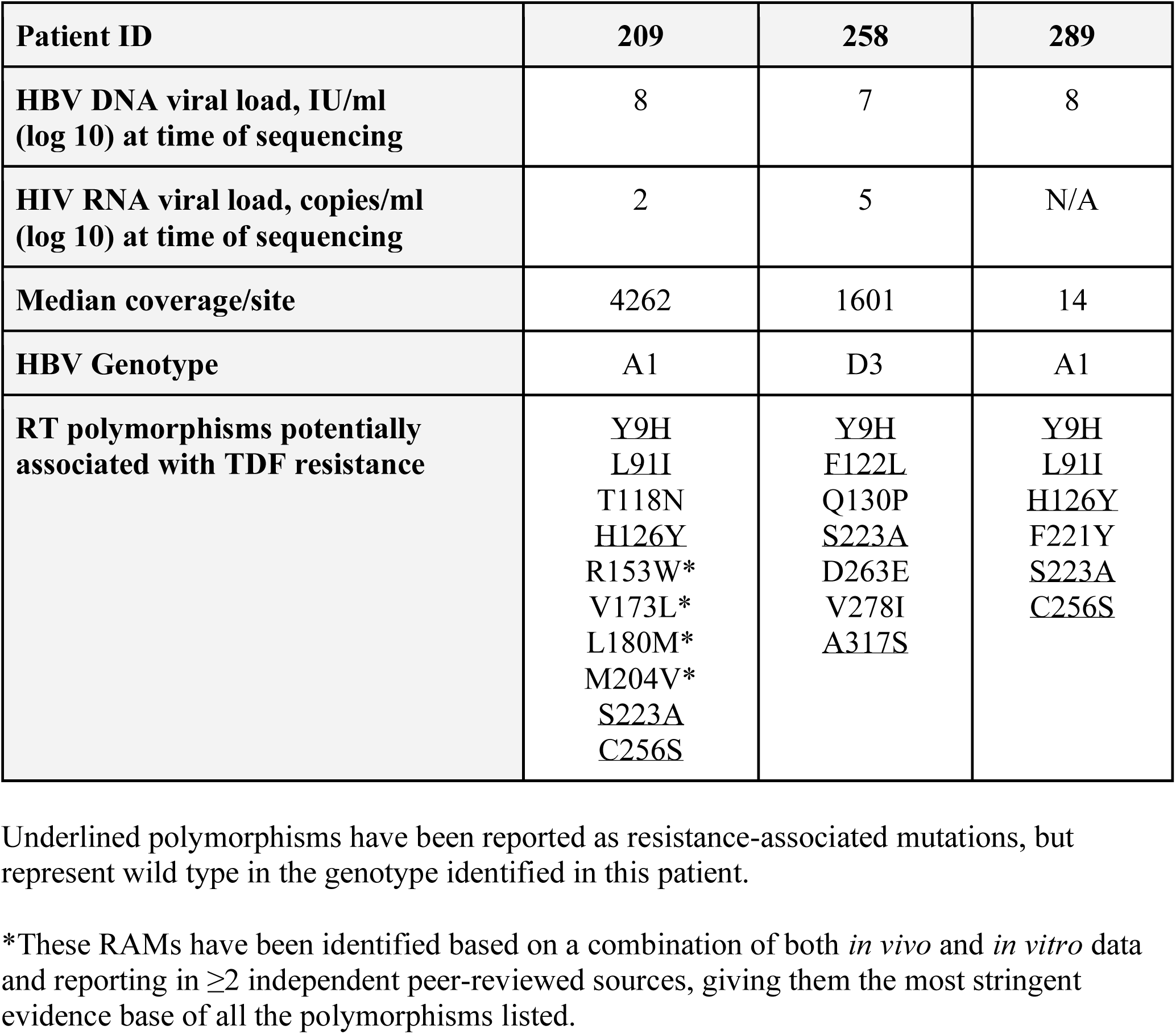
Summary of HBV sequence data generated from three South African adults with persistent HBV viraemia ≥20,000 IU/ml despite >12 months prescription of TDF therapy. Patients 209 and 258 had HIV co-infection; patient 289 was HIV negative.

In HBV sequences from patient 209, we observed ten potential TDF RAMs. Among these, four polymorphisms, (R153W, V173L, L180M, and M204V) have been identified based on the most stringent combination of evidence, reflecting both *in vivo* and *in vitro* data, and reported in ≥2 independent peer-reviewed sources (6). While M204I/V and L180M are best recognised in association with 3TC resistance (21), they may also be part of a combination of polymorphisms that underpins TDF resistance. In patients 258 and 289, seven and five putative TDF RAMs were observed, respectively. At a deep sequence level, the RAMs we observed in all three patients were present in >95% of HBV reads.

Together, these observations are in keeping with established evidence that suggests a high genetic barrier to TDF resistance, requiring the selection of suites of multiple mutations to mediate a drug-resistant phenotype.

RAMs T118C/G, F122L and Q130S have been previously identified in HBV sequences obtained from individuals in whom viraemia was not suppressed by TDF (4,22)). Interestingly, we identified different substitutions at these three sites: T118N and F122Y (in patient 209) and Q130P (in patient 258); Table 1. The amino acid substitutions we observed have similar chemical properties to those previously reported (i.e non-polar side-chain and neutral pH) and therefore might have a similar effects in terms of mediating drug resistance.

For each potential RAM identified in our HBV sequences we reviewed the prevalence of the variants in all available published HBV sequences (Table 2). In all three cases, a number of polymorphisms that have been reported as TDF RAMs actually represent the most common residue in this genotype (based on reference sequences A1 (FJ692557) and D3 (FJ692507) (23)). These sites are indicated by underlining in Table 1. Of note, for patient 289, four of five TDF RAMs actually occur at a consensus level.

**Table 2:** **Frequency of putative TDF RAMs in A (n=840) and D (n=958) sequences downloaded from Hepatitis B Virus Database (HBVdb) - https://hbvdb.ibcp.fr/HBVdb/ on the 20th of November 2018**.

| **Polymorphism in HBV RT** | **Frequency in genotype A sequences** | **Frequency in genotype D sequences** |
| --- | --- | --- |
| Y9H | 286/ 290 (98.6%) | 556/566 (98.2%) |
| L91I | 290/290 (100%) | 41/566 (7.2%) |
| T118N | 283/290 (97.6%) | 542/566 (95.8%) |
| F122Y/L | 8/290 (2.8%) | 82/566 (14.5%) |
| H/126Y | 225/290 (77.6%) | 4/566 (0.7%) |
| Q130P | 1/290 (0.3%) | 77/566 (13.6%) |
| R153W | 228/290 (78.6%) | 22/566 (3.9%) |
| V173L | 0/290 (0%) | 4/566 (0.7%) |
| L180M | 7/290 (2.4%) | 13/566 (2.3%) |
| M204I/V | 7/290 (2.4%) | 25/566 (4.4%) |
| F221Y | 282/290 (97.2%) | 28/566 (4.9%) |
| S223A | 289/290 (99.7%) | 563/566 (99.5%) |
| C256S | 279/290 (96.2%) | 65/566 (11.5%) |
| D263E | 1/290 (0.3%) | 111/566 (19.6%) |
| V278I | 16/290 (5.5%) | 55/566 (9.7%) |
| A317S | 4/290 (1.4%) | 531/566 (93.8%) |

## DISCUSSION

### Summary of findings

We here present the first evidence of potential TDF resistance in adults being treated for chronic HBV in Africa, combining phenotypic observations of a TDF resistant phenotype with viral sequencing data. In particular, case 209 represents strong evidence for HBV resistance to TDF, on the basis of high HBV DNA viral load (>10^8^ IU/ml) despite 60 months of therapy, with a suppressed HIV viral load, and a combination of sequence polymorphisms that have been reported in association with TDF resistance. The other two cases illustrate the potential difficulties in discriminating between drug resistance and incomplete adherence, and may also reflect the long timelines sometimes associated with viraemic suppression on TDF (20). Overall, our observations add to the emerging global evidence of TDF resistance and reinforce the need for therapeutic advances for HBV, including novel agents or drug combinations, to support global progress towards eliminating viral hepatitis as a public health threat by the year 2030.

### Significance of resistance data for Africa

We have previously highlighted the paucity of HBV drug resistance data representing Africa (8). While we here report a picture in keeping with definitive TDF resistance in just one individual, collection of the clinical, laboratory and molecular data that are required to identify such cases is rare in most settings across Africa (24). As TDF is widely used in Africa as a component of first line ART, it is possible that the selection of resistance mutations in HBV is more prevalent than currently reported (8,14). Engagement and investment in this important question are urgently required to further enable us to determine the prevalence and clinical significance of TDF RAMs in different African populations.

### Interpretation of sequence data

In participant 209, five of ten polymorphisms (H126Y, R153W, V173L, L180M and M204V) have been assessed for resistance through sequencing of isolates obtained from individuals in whom viraemia was not suppressed by TDF, and subsequently validated *in vitro* (6,8), representing the most robust evidence for a clinically significant role in drug resistance. RAMs V173L, L180M and M204V are known to cause HBV resistance to 3TC. Given that this participant was on both TDF and 3TC, it is likely that exposure to 3TC resulted in the development of RAMs V173L, L180M and M204V, possibly setting the scene for the subsequent development of cross-resistance to TDF. While TDF is considered effective even with prior use of other NAs (25), there is evidence to show a decrease in effectiveness of TDF in HBV viral suppression among individuals with prolonged 3TC exposure (26).

In participant 258, seven RAMs were identified that have also been reported from other individuals in whom HBV viraemia was not suppressed on TDF (6). While we identified five potential RAMs in participant 289, viraemia was eventually suppressed after 30 months of treatment, raising a question as to whether genuinely TDF resistance is present. Furthermore, all amino acid variants in this sequence occur frequently in genotype A sequences, making it less likely that drug resistant polymorphisms have been selected *de novo*. Interestingly, the large flares in liver enzymes in this individual (Fig 2C) may represent evidence of a host immunological response in parallel with drug therapy accounting for the reduction in HBV DNA viraemia.

### Role of combination antiviral therapy for HBV

TDF and FTC and/or 3TC are part of the first line treatment regimen for HIV infection (14). Although FTC is not part of the recommended treatment options for chronic HBV infection, it has been shown to have activity against HBV, and a combination of TDF and FTC has been shown to have better viral suppression compared to TDF monotherapy among treatment naive individuals with chronic HBV infection (27). However, another study reported no difference in effect between TDF monotherapy versus combined therapy of TDF and 3TC/FTC in managing chronic HBV infection among HBV monoinfected individuals (28). Among HIV/HBV coinfected individuals, a combination of 3TC and TDF is considered more efficacious compared to 3TC monotherapy among HIV/HBV coinfected individuals (29).

### Caveats and limitations

The data we present here are limited to only three individuals, with different clinical phenotypes (Fig 2). In patient 289, despite a high viral load we achieved a limited sequencing coverage depth (Table 2), which reduces the sensitivity for detection of mutations occurring as minority variants. Incomplete treatment adherence can also result in virological breakthrough, and assessment of drug levels in plasma and/ or PBMCs is required to provide more objective information on treatment exposure (e.g. (30)), especially in HBV monoinfected patients in whom suppression of HIV cannot be used as a surrogate measure of treatment compliance.

To date, there are limited published data in support of TDF resistance, and the precise contribution of specific polymorphisms, either alone or acting in collaboration, remains to be determined with certainty. There is heterogeneity in approaches used to identify and describe mutations associated with TDF resistance; studies reporting these mutations apply differing approaches to defining TDF resistance (6). Furthermore, some polymorphisms described in association with TDF resistance represent the wild type sequence in certain genotypes. This phenomenon has not been discussed in detail for HBV infection, but is well described for hepatitis C virus (HCV) infection, in which certain subgenotypes are more treatment-resistant because RAMs are present in the wild type sequence (31,32). To further our understanding of this effect in HBV, there is a pressing need for more clinical data regarding patients in whom TDF does not mediate viraemic suppression, combined with *in vitro* data from assays that can confirm a resistant phenotype and determine the impact of mutations acting alone and together. To date, the crystal structure of HBV RT has not been solved, but advances in structural biology may also be important to provide mechanistic insights.

### Summary

Our findings add to the evidence that mutations in HBV RT can underpin a TDF resistant phenotype. This is the first time these RAMs have been reported from Africa supported by clinical evidence of TDF resistance. In HBV/HIV coinfected patients, persistent HBV viraemia in the context of HIV suppression could be regarded as an indicator or ‘red flag’ for TDF resistance. More data are required to determine the frequency and clinical impact of RAMs, to investigate the role of combination therapy, and to support the development of new antiviral agents.

## Data Availability

HBV sequence data are available at GenBank [accession numbers MT210032, MT210033, MT210034]

## Data availability

HBV sequence data are available at GenBank [accession numbers MT210032, MT210033, MT210034]

## Authors Contribution Statement

JM, TM and PM conceived the study. TM and PM acquired funding and ethics approval. TM, MvS, SH, JT, WP, CvR recruited patients and collected clinical data. JM, AM, DB and MdC generated sequence data. JM, TM, AM, JS, VS, MC and PM analysed the data. JM, AM and PM wrote the manuscript. All authors revised and approved the final manuscript.

---

**Supplementary Table 1:**
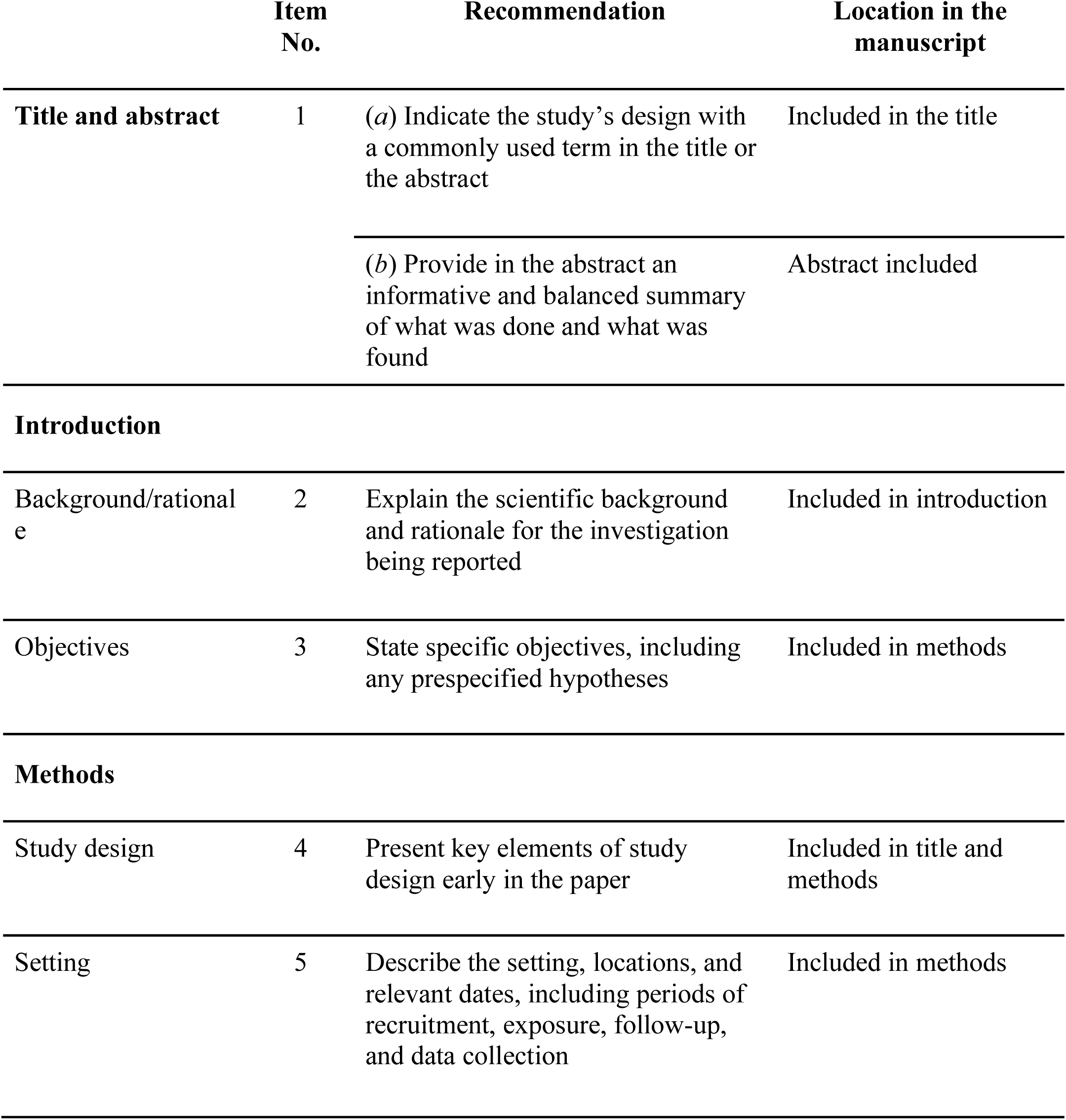
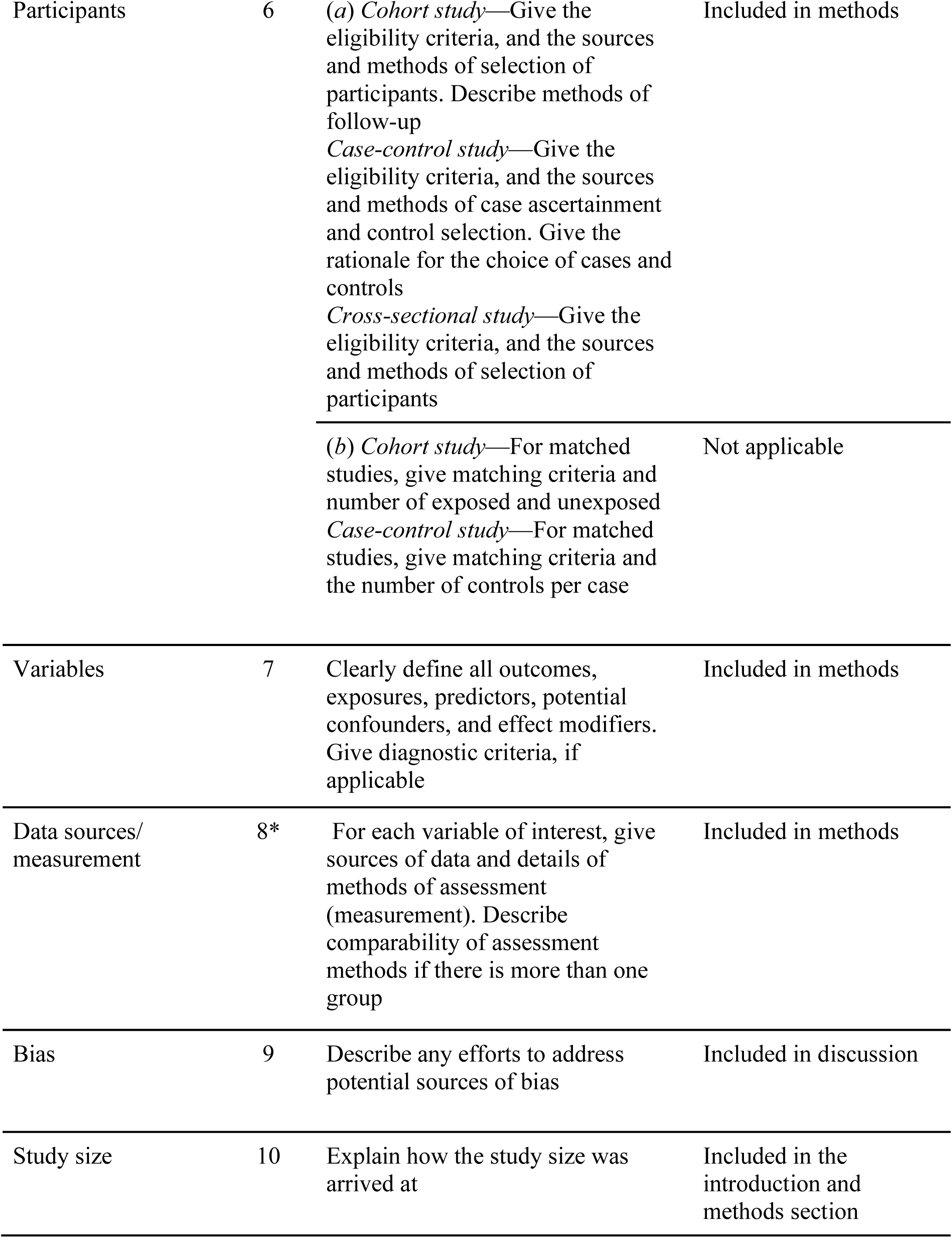
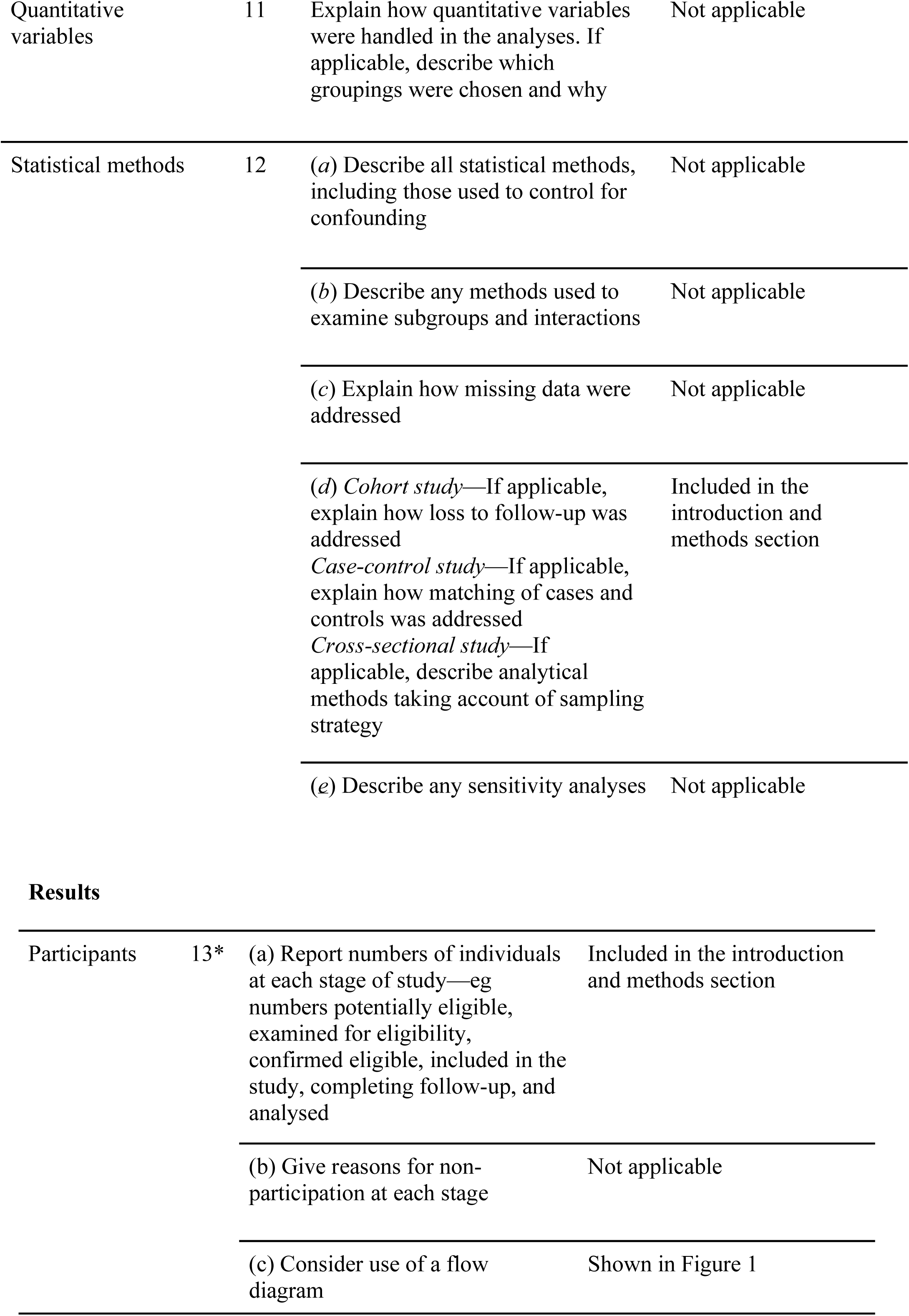
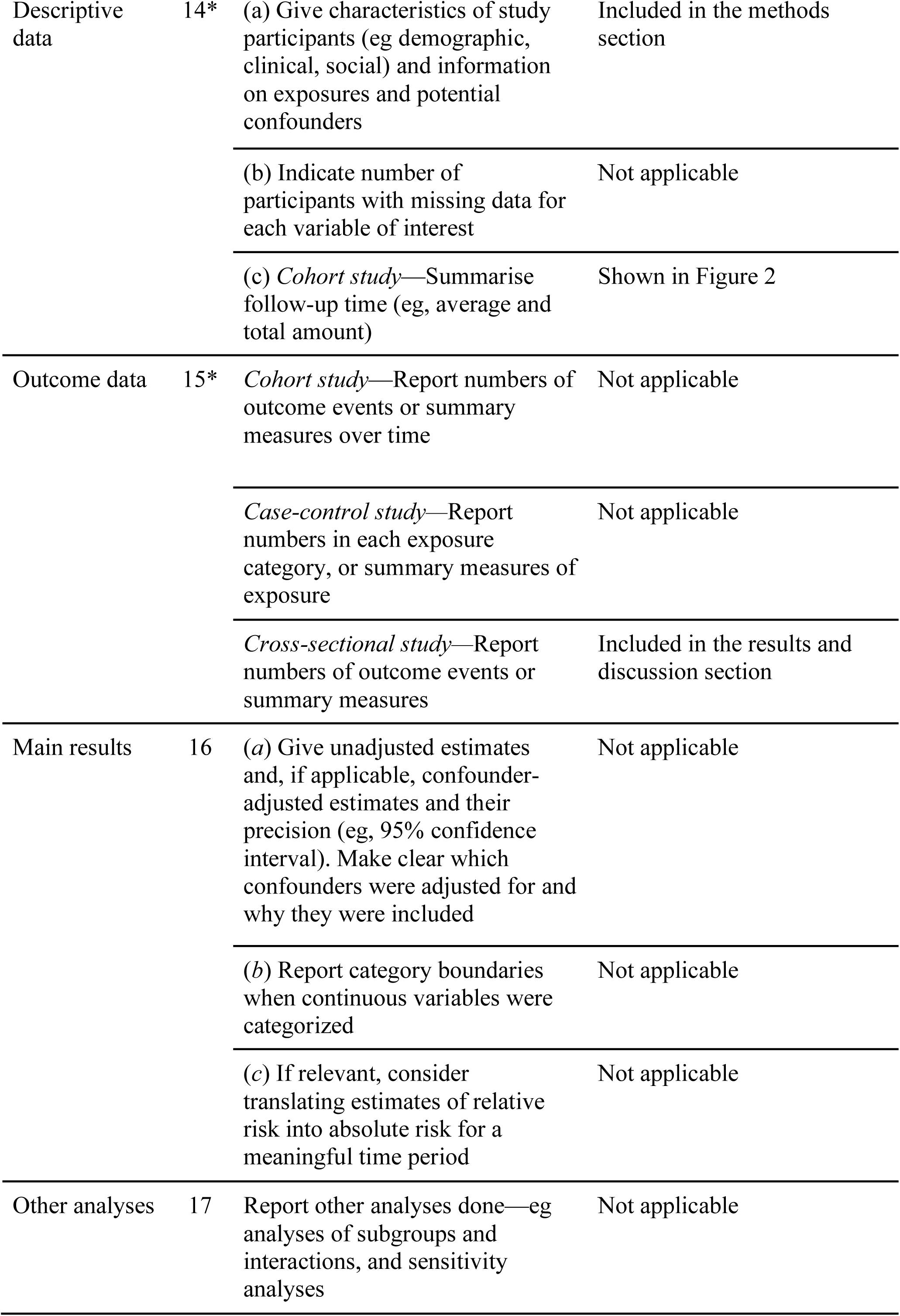
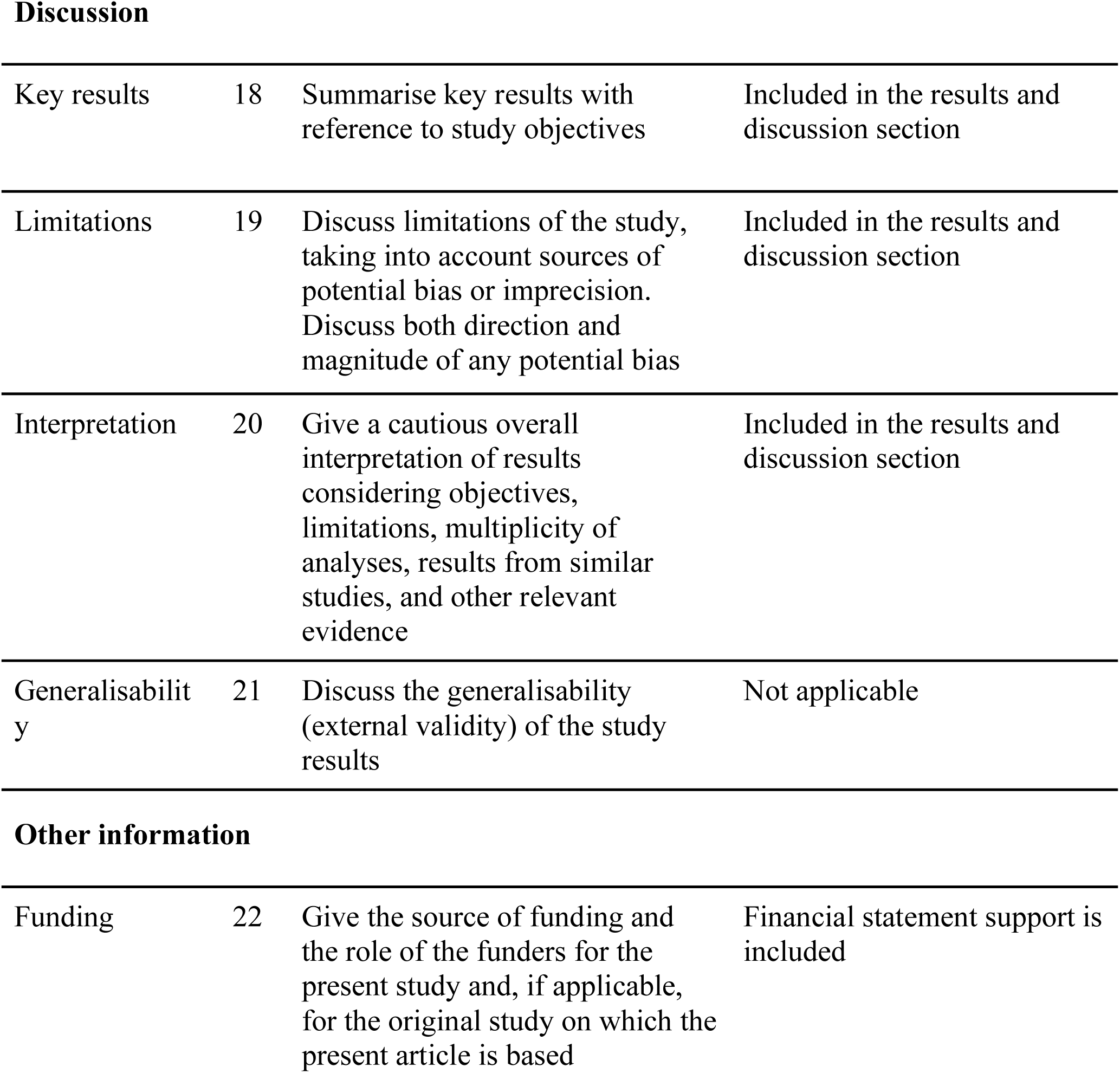
STROBE Statement

|  | Item No. | Recommendation | Location in the manuscript |
| --- | --- | --- | --- |
| Title and abstract | 1 | (a) Indicate the study’s design with a commonly used term in the title or the abstract | Included in the title |
|  |  | (b) Provide in the abstract an informative and balanced summary of what was done and what was found | Abstract included |
| Introduction |  |  |  |
| Background/rationale | 2 | Explain the scientific background and rationale for the investigation being reported | Included in introduction |
| Objectives | 3 | State specific objectives, including any prespecified hypotheses | Included in methods |
| Methods |  |  |  |
| Study design | 4 | Present key elements of study design early in the paper | Included in title and methods |
| Setting | 5 | Describe the setting, locations, and relevant dates, including periods of recruitment, exposure, follow-up, and data collection | Included in methods |
| Participants | 6 | (a) *Cohort study*—Give the eligibility criteria, and the sources and methods of selection of participants. Describe methods of follow-up<br>*Case-control study*—Give the eligibility criteria, and the sources and methods of case ascertainment and control selection. Give the rationale for the choice of cases and controls<br>*Cross-sectional study*—Give the eligibility criteria, and the sources and methods of selection of participants | Included in methods |
|  |  | (b) *Cohort study*—For matched studies, give matching criteria and number of exposed and unexposed<br>*Case-control study*—For matched studies, give matching criteria and the number of controls per case | Not applicable |
| Variables | 7 | Clearly define all outcomes, exposures, predictors, potential confounders, and effect modifiers. Give diagnostic criteria, if applicable | Included in methods |
| Data sources/<br>measurement | 8* | For each variable of interest, give sources of data and details of methods of assessment (measurement). Describe comparability of assessment methods if there is more than one group | Included in methods |
| Bias | 9 | Describe any efforts to address potential sources of bias | Included in discussion |
| Study size | 10 | Explain how the study size was arrived at | Included in the introduction and methods section |
| Quantitative variables | 11 | Explain how quantitative variables were handled in the analyses. If applicable, describe which groupings were chosen and why | Not applicable |
| Statistical methods | 12 | (a) Describe all statistical methods, including those used to control for confounding | Not applicable |
|  |  | (b) Describe any methods used to examine subgroups and interactions | Not applicable |
|  |  | (c) Explain how missing data were addressed | Not applicable |
|  |  | (d) *Cohort study*—If applicable, explain how loss to follow-up was addressed<br>*Case-control study*—If applicable, explain how matching of cases and controls was addressed<br>*Cross-sectional study*—If applicable, describe analytical methods taking account of sampling strategy | Included in the introduction and methods section |
|  |  | (e) Describe any sensitivity analyses | Not applicable |
| Participants | 13* | (a) Report numbers of individuals at each stage of study—eg numbers potentially eligible, examined for eligibility, confirmed eligible, included in the study, completing follow-up, and analysed | Included in the introduction and methods section |
|  |  | (b) Give reasons for non-participation at each stage | Not applicable |
|  |  | (c) Consider use of a flow diagram | Shown in Figure 1 |
| Descriptive data | 14* | (a) Give characteristics of study participants (eg demographic, clinical, social) and information on exposures and potential confounders | Included in the methods section |
|  |  | (b) Indicate number of participants with missing data for each variable of interest | Not applicable |
|  |  | (c) *Cohort study*—Summarise follow-up time (eg, average and total amount) | Shown in Figure 2 |
| Outcome data | 15* | *Cohort study*—Report numbers of outcome events or summary measures over time | Not applicable |
|  |  | *Case-control study*—Report numbers in each exposure category, or summary measures of exposure | Not applicable |
|  |  | *Cross-sectional study*—Report numbers of outcome events or summary measures | Included in the results and discussion section |
| Main results | 16 | (a) Give unadjusted estimates and, if applicable, confounder-adjusted estimates and their precision (eg, 95% confidence interval). Make clear which confounders were adjusted for and why they were included | Not applicable |
|  |  | (b) Report category boundaries when continuous variables were categorized | Not applicable |
|  |  | (c) If relevant, consider translating estimates of relative risk into absolute risk for a meaningful time period | Not applicable |
| Other analyses | 17 | Report other analyses done—eg analyses of subgroups and interactions, and sensitivity analyses | Not applicable |
| Key results | 18 | Summarise key results with reference to study objectives | Included in the results and discussion section |
| Limitations | 19 | Discuss limitations of the study, taking into account sources of potential bias or imprecision. Discuss both direction and magnitude of any potential bias | Included in the results and discussion section |
| Interpretation | 20 | Give a cautious overall interpretation of results considering objectives, limitations, multiplicity of analyses, results from similar studies, and other relevant evidence | Included in the results and discussion section |
| Generalisability | 21 | Discuss the generalisability (external validity) of the study results | Not applicable |
| Other information |  |  |  |
| Funding | 22 | Give the source of funding and the role of the funders for the present study and, if applicable, for the original study on which the present article is based | Financial statement support is included |

## REFERENCES

1. World Health Organisation. (2019). Guidelines for the prevention, care and treatment of persons with chronic hepatitis B infection. Available at: http://www.who.int/hepatitis/publications/hepatitis-b-guidelines/en/ [Accessed February 22, 2020]

2. Marcellin P, Wong DK, Sievert W, Buggisch P, Petersen J, Flisiak R, Manns M, Kaita K, Krastev Z, Lee SS, et al. (2019). Ten-year efficacy and safety of tenofovir disoproxil fumarate treatment for chronic hepatitis B virus infection. Liver Int. 39 :1868–1875.

3. Mokaya J, Burn EAO, Tamandjou CR, Goedhals D, Barnes EJ, Andersson M, Pinedo-Villanueva R, Matthews PC. (2019). Modelling cost-effectiveness of tenofovir for prevention of mother to child transmission of hepatitis B virus (HBV) infection in South Africa. BMC Public Health. 19 :829.

4. Cho WH, Lee HJ, Bang KB, Kim SB, Song IH. (2018). Development of tenofovir disoproxil fumarate resistance after complete viral suppression in a patient with treatment-naïve chronic hepatitis B: A case report and review of the literature. World J Gastroenterol. 24 :1919–1924.

5. Park E-S, Lee AR, Kim DH, Lee J-H, Yoo J-J, Ahn SH, Sim H, Park S, Kang HS, Won J, et al. (2019). Identification of a quadruple mutation that confers tenofovir resistance in chronic hepatitis B patients. J Hepatol. 70 :1093–1102.

6. Mokaya J, McNaughton AL, Bester PA, Goedhals D, Barnes E, Marsden BD, Matthews PC. (2010). Hepatitis B virus resistance to tenofovir: fact or fiction? A synthesis of the evidence to date. MedRxiv [preprint]. Available at: https://www.medrxiv.org/content/10.1101/19009563v1 [Accessed February 22, 2020]

7. Lemoine M, Eholié S, Lacombe K. (2015). Reducing the neglected burden of viral hepatitis in Africa: strategies for a global approach. J Hepatol. 62 :469–476.

8. Mokaya J, McNaughton AL, Hadley MJ, Beloukas A, Geretti A-M, Goedhals D, Matthews PC. (2018). A systematic review of hepatitis B virus (HBV) drug and vaccine escape mutations in Africa: A call for urgent action. PLOS Neglected Tropical Diseases. 12 :e0006629. doi: 10.1371/journal.pntd.0006629

9. World Health Organisation. Global hepatitis report. (2017). Available at: https://www.who.int/hepatitis/publications/global-hepatitis-report2017/en/ [Accessed February 22, 2020]

10. World Health Organisation. (2016). Combating Hepatitis B and C to reach elimination by 2030. Available at: http://apps.who.int/iris/bitstream/10665/206453/1/WHO_HIV_2016.04_eng.pdf?ua=1 [Accessed February 22, 2020]

11. Cooke GS, Andrieux-Meyer I, Applegate TL, Atun R, Burry JR, Cheinquer H, Dusheiko G, Feld JJ, Gore C, Griswold MG, et al. (2019). Accelerating the elimination of viral hepatitis: a Lancet Gastroenterology & Hepatology Commission. Lancet Gastroenterol Hepatol 4 :135–184.

12. Maponga TG, McNaughton AL, Van Schalkwyk M, Hugo S, Nwankwo C, Taljaard J, Mokaya J, Smith DA, van Vuuren C, Goedhals D, et al. (2019). Treatment advantage in HBV/HIV coinfection compared to HBV monoinfection in a South African cohort. MedRxiv [preprint]. Available at: https://www.medrxiv.org/content/10.1101/19007963v2 [Accessed February 22, 2020]

13. Spearman CWN, Sonderup MW, Botha JF, van der Merwe SW, Song E, Kassianides C, Newton KA, Hairwadzi HN, Division of Hepatology, Department of Medicine, University of Cape Town, South Africa. (2013). South African guideline for the management of chronic hepatitis B: 2013. S Afr Med J. 103 :337–349.

14. World Health Organisation. (2019). Updated recommendations on first-line and second-line antiretroviral regimens and post-exposure prophylaxis and recommendations on early infant diagnosis of HIV. Available at: http://www.who.int/hiv/pub/guidelines/ARV2018update/en/ [Accessed March 9, 2020]

15. HIV/AIDS Treatment Guidelines. AIDSinfo Available at: https://aidsinfo.nih.gov/guidelines [Accessed March 12, 2020]

16. Meintjes G, Moorhouse MA, Carmona S, Davies N, Dlamini S, van Vuuren C, Manzini T, Mathe M, Moosa Y, Nash J, et al. (2017). Adult antiretroviral therapy guidelines 2017. South Afr J HIV Med. 18 :776.

17. McNaughton AL, Roberts HE, Bonsall D, de Cesare M, Mokaya J, Lumley SF, Golubchik T, Piazza P, Martin JB, de Lara C, et al. (2019). Illumina and Nanopore methods for whole genome sequencing of hepatitis B virus (HBV). Sci Rep. 9 :7081.

18. Martel N, Gomes SA, Chemin I, Trépo C, Kay A. (2013). Improved rolling circle amplification (RCA) of hepatitis B virus (HBV) relaxed-circular serum DNA (RC- DNA). J Virol Methods 193 :653–659.

19. Singer JB, Thomson EC, McLauchlan J, Hughes J, Gifford RJ. (2018). GLUE: A flexible software system for virus sequence data. BMC Bioinformatics 19, 532

20. Min IS, Lee CH, Shin IS, Lee NE, Son HS, Kim SB, Seo SY, Kim SH, Kim SW, Lee SO, et al. (2019). Treatment Outcome and Renal Safety of 3-Year Tenofovir Disoproxil Fumarate Therapy in Chronic Hepatitis B Patients with Preserved Glomerular Filtration Rate. Gut Liver. 13 :93–103.

21. Beloukas A, Geretti AM. (2017). Hepatitis B Virus Drug Resistance. In: Antimicrobial Drug Resistance. Cham: Springer International Publishing.

22. Mikulska M, Taramasso L, Giacobbe DR, Caligiuri P, Bruzzone B, Di Biagio A, Viscoli C. (2012). Case report: Management and HBV sequencing in a patient co-infected with HBV and HIV failing tenofovir. Journal of Medical Virology 84 :1340–1343. doi: 10.1002/jmv.23338

23. McNaughton AL, Revill PA, Littlejohn M, Matthews PC, Ansari MA. (2020). Analysis of genomic-length HBV sequences to determine genotype and subgenotype reference sequences. J Gen Virol doi: 10.1099/jgv.0.001387

24. O’Hara GA, McNaughton AL, Maponga T, Jooste P, Ocama P, Chilengi R, Mokaya J, Liyayi MI, Wachira T, Gikungi DM, et al. (2017). Hepatitis B virus infection as a neglected tropical disease. PLoS Negl Trop Dis. 11 :e0005842.

25. Price H, Dunn D, Pillay D, Bani-Sadr F, de Vries-Sluijs T, Jain MK, Kuzushita N, Mauss S, Núñez M, Nüesch R, et al. (2013). Suppression of HBV by tenofovir in HBV/HIV coinfected patients: a systematic review and meta-analysis. PLoS One 8 :e68152.

26. Kim HN, Nina Kim H, Rodriguez CV, Van Rompaey S, Eron JJ, Thio CL, Crane HM, Overton ET, Saag MS, Martin J, et al. (2014). Factors Associated With Delayed Hepatitis B Viral Suppression on Tenofovir Among Patients Coinfected With HBV-HIV in the CNICS Cohort. JAIDS Journal of Acquired Immune Deficiency Syndromes. 66 :96–101.

27. Cui G, Xu X, Diao H. (2015). Comparative Meta-Analysis of Tenofovir Disoproxil Fumarate versus Emtricitabine and Tenofovir Disoproxil Fumarate as Treatments for Patients with Chronic Hepatitis B. Sci Rep 5 :1–11.

28. Berg T, Marcellin P, Zoulim F, Moller B, Trinh H, Chan S, Suarez E, Lavocat F, Snow–Lampart A, Frederick D, et al. (2010). Tenofovir Is Effective Alone or With Emtricitabine in Adefovir-Treated Patients With Chronic-Hepatitis B Virus Infection. Gastroenterology. 139 :1207–1217.e3.

29. Luo A, Jiang X, Ren H. (2018). Lamivudine plus tenofovir combination therapy versus lamivudine monotherapy for HBV/HIV coinfection: a meta-analysis. Virol J. 15 :139.

30. Louissaint NA, Cao Y-J, Skipper PL, Liberman RG, Tannenbaum SR, Nimmagadda S, Anderson JR, Everts S, Bakshi R, Fuchs EJ, et al. (2013). Single dose pharmacokinetics of oral tenofovir in plasma, peripheral blood mononuclear cells, colonic tissue, and vaginal tissue. AIDS Res Hum Retroviruses. 29 :1443–1450.

31. Davis C, Mgomella GS, da Silva Filipe A, Frost EH, Giroux G, Hughes J, Hogan C, Kaleebu P, Asiki G, McLauchlan J, et al. (2019). Highly Diverse Hepatitis C Strains Detected in Sub-Saharan Africa Have Unknown Susceptibility to Direct-Acting Antiviral Treatments. Hepatology. 69 :1426–1441.

32. Fourati S, Rodriguez C, Hézode C, Soulier A, Ruiz I, Poiteau L, Chevaliez S, Pawlotsky J-M. (2019). Frequent Antiviral Treatment Failures in Patients Infected With Hepatitis C Virus Genotype 4, Subtype 4r. Hepatology. 69 :513–523.

